# Music Therapy in Pediatric and Neonatal Intensive Care Units: A Systematic Review and Meta-Analysis

**DOI:** 10.1101/2024.10.13.24314254

**Authors:** Luiza Lorenz Cavalcante, Adília Maria Pires Sciarra, Moacir Fernandes de Godoy

## Abstract

**Introduction:** Music Therapy is a form of therapy that involves the musical sphere to promote the well-being and improvement of patients’ vital conditions. Conversely, intensive care units are normally recognized as stressful and threatening environments. Therefore, it can be used to contribute to maintaining the mental health of these patients.

**Method:** A systematic review with meta-analysis of Music Therapy in pediatric and neonatal intensive care units was performed. The methodology followed the PRISMA 2020 guidelines, with 10 articles analyzed, and a total of approximately 737 patients. The vital signs assessed were respiratory rate, heart rate, oxygen saturation, blood pressure, and perceived pain intensity. Additionally, a comparison was made between different types of music for each population studied.

**Results:** It was observed that Music Therapy reduced the respiratory rate and pain sensitivity index in the pediatric ICU by 1.92 bpm (p-value: 0.004) and 2.09 Wong-Baker FACES units (p-value: <0.0001), respectively. Additionally, it has increased oxygen saturation in pediatric and neonatal ICUs by 0.37 (p-value: 0.001) and 0.2 (p-value: 0.04), respectively. The favorite kind of music listened to by the patients presented better results than the others.

**Conclusion:** There is a scarcity of articles that would allow a comprehensive analysis of this theme, specifically studying different types of this therapy with these populations. Additionally, based on the results presented in the study, it is evident that music therapy is a potential treatment that can be utilized in a hospital setting. Few articles facilitate a comprehensive analysis of this theme, particularly regarding various types of therapy for these populations. Furthermore, the study’s results have pointed out that music therapy is a viable treatment option in a hospital setting.

## Introduction

Music Therapy is a therapeutic approach that utilizes the power of music to enhance the well-being and overall health of patients. It aims to reduce respiratory rate and improve mental health through individual or group sessions that incorporate musical instruments, singing, melodies, and recordings.

This type of treatment is widely recognized in various countries across Oceania, Europe, Asia, and the Americas. Consequently, organizations have been established to supervise, train, and assist other health institutions to enhance their effectiveness in patient care. Other organizations, such as the American Music Therapy Association and the European Music Therapy Confederation, provide a list of active groups in several countries, including the United States, Australia, Finland, Germany, and New Zealand. (1,2).

The UBAM, a renowned organization in Brazil, exemplifies the significance of music therapy in the Brazilian healthcare system. Furthermore, the government recognized this type of treatment as an integrative and complementary practice within the Unified Health System (SUS) on March 27, 2017 (3)

It is important to emphasize that music therapy focuses on maintaining the mental health integrity of patients during stressful activities and improving some physical aspects. It can be regarded as a treatment for Post-Intensive Care Syndrome (PICS), a mental illness that affects numerous patients in Intensive Care Units. They usually develop emotional disorders, which can lead to severe depression and a poor quality of life.

Data analysis reveals that numerous current studies demonstrate the efficacy of music therapy, especially for children in the intensive care unit. It also indicates that there are various types of approaches and modalities that professionals can use in this therapy. From this perspective, there is a clear need to analyze the applicability of music therapy in the ICU. Furthermore, it is imperative to assess the most efficacious intervention among the numerous options available in Music Therapy.

### Objectives

The central objective of this systematic review was to analyze the influence on vital parameters (heart rate, respiratory rate, blood pressure, oxygen saturation, and pain sensation level) most highlighted among the articles about the use of Music Therapy in pediatric and neonatal ICU, and to determine the specific objective, the authors evaluated the different outcomes according to the type of management of this treatment, since music therapy is present in the medical field with diverse instruments and musical styles.

## Method

Several scientific articles published in different databases were investigated. It is worth mentioning that since this is a systematic literature review, the researched documents were analyzed objectively and impartially, during the period from July to September 2023. Several revisions of the values and articles used throughout the research were carried out, although they were done manually by the authors.

The analysis was based on understanding the methodology employed, the outcomes obtained, and the conclusions drawn by the authors. Furthermore, the data from the period of 2013 to 2023 was considered as the publication date.

The access to the chosen scientific literature database for the survey was provided through an institutional subscription: “Faculdade de Medicina de São José do Rio Preto”, SP, Brazil; “Portal de Periódicos Capes” (CAPES MEC); remote access. It was provided free access to search publications, which meant that they were available to the researchers.

It is worth noting that databases available on internationally recognized platforms for scientific research, such as Medline, LILACS, and PubMed, were also used. The researchers were able to access all of them due to their availability in the public domain. However, not all studies found in these databases were used because some did not allow their reading to be entirely free. Following Article VI of Resolution No. 510/16, Subjects (CEP/FAMERP) were not necessary.

Finally, the submission of this study to the Research Ethics Committee with Human (2016) of the Ministry of Health of Brazil, National Health Council, establishes that research using publicly available information, such as this study, should not be evaluated or registered by the CEP/CONEP system. PRISMA (2020) methodology (12) was used to analyze and filter the database used for the systematic review and meta-analysis. Therefore, the search for articles was based on criteria and flowcharts pre-established by this methodological system.

In this manner, combinations of keywords were developed to filter the works in different research databases. They are listed below.

### Medline

● (ti: music therapy”) AND (ti: intensive care”)) AND (children) AND NOT (neurodevelopment) AND NOT (newborn) AND NOT (neonatal)
● (ti:(”music therapy”)) AND (ti:(”intensive care”)) AND (neonatal) AND NOT (neurodevelopment) AND NOT (children)
● (ti:(”music therapy”)) AND (ti:(”intensive care”)) AND (newborn) AND NOT (neurodevelopment) AND NOT (children)
● (ti:(”music therapy”)) AND (ti:(”intensive care”)) AND (infant) AND NOT (neurodevelopment) AND NOT (children)
● (ti:(music)) AND (ti:(”intensive care”)) AND (infant) AND NOT (neurodevelopment) AND NOT (children)
● (ti:(music)) AND (ti:(”intensive care”)) AND (neonatal) AND NOT (neurodevelopment) AND NOT (children)
● (ti:(music)) AND (ti:(”intensive care”)) AND (infant) AND NOT (neurodevelopment) AND NOT (children)
● (ti:(music)) AND (ti:(”intensive care”)) AND (newborn) AND NOT (neurodevelopment) AND NOT (children)

### LILACS

● “music therapy” “intensive care” children
● “music therapy” “intensive care infant newborn neonatal
● music “intensive care” infant newborn neonatal

### PubMed

● “music therapy” AND “intensive care” AND children NOT neurodevelopment NOT newborn NOT neonatal
● “music therapy” AND “intensive care” AND newborn NOT neurodevelopment NOT children
● “music therapy” AND “intensive care” AND neonatal NOT neurodevelopment NOT children
● “music therapy” AND “intensive care” AND infant NOT neurodevelopment NOT children
● music “intensive care” infant NOT neurodevelopment NOT children
● music “intensive care” newborn NOT neurodevelopment NOT children
● music “intensive care” neonatal NOT neurodevelopment NOT children

### Web of Science

● “music therapy” “intensive care” children -neurodevelopment -newborn -neonatal
● “music therapy” “intensive care” children
● “music therapy””intensive care” neonatal NOT neurodevelopment NOT children
● “music therapy””intensive care” newborn NOT neurodevelopment NOT children
● “music therapy””intensive care” infant NOT neurodevelopment NOT children
● music “intensive care” infant NOT neurodevelopment NOT children
● music “intensive care” neonatal NOT neurodevelopment NOT children
● music “intensive care” newborn NOT neurodevelopment NOT children

Furthermore, articles that were beyond the predetermined analysis period (2013-2023) were disregarded. As a result, it was possible to determine an absolute quantity of articles; 595 studies (141 for children and 454, for neonatal) that would later be analyzed and filtered.

The initial filtering procedure consisted of eliminating duplicate studies, specifically 371 studies that had appeared on multiple occasions in other research databases or with a different keyword sequence. Furthermore, 107 articles were excluded because their titles contained information that was outside the analyzed repertoire. For example, if the study was different from a quasi-experimental study or a randomized controlled clinical trial, or if they emphasized professionals’ perceptions regarding Music Therapy, among other reasons.

The subsequent filtering process comprised an analysis of the title and abstract of the works, with only those that were closest to the inclusion criteria being considered for inclusion in the subsequent review process. At the end of the process, 40 studies were excluded, and 57 articles were passed.

The inclusion and exclusion criteria are the following:

### Included

Articles with significant keywords in the title that align with the research theme, such as “music therapy,” “intensive care,” CICU, NICU, SICU, among others; publications published during the predetermined period (2013-2023); obtaining the essential data from both the experimental and control groups, including at least one of the following parameters: heart rate, respiratory rate, blood pressure (systolic and diastolic), oxygen saturation, and patient pain sensitivity level.

### Excluded

Articles with titles containing keywords outside the research theme, such as “autism” and “neurodevelopment”; documents of a different type than quasi-experimental study, randomized controlled clinical trial, and controlled clinical trial; those without permitted access; texts addressing a different age group than the one evaluated; when the analyzed group had a wide range of vital signs due to differing ages. If the article showed discrepancies in its information, and when the article exhibits discreetness.

During the third filtering stage, articles that could not be accessed in full text were disregarded because they were not in the public domain or saved in the research database at the time of this study’s development. Under these conditions, sixteen articles were excluded.

The final step comprised a thorough reading of the remaining texts, with only those that met the inclusion criteria included in the study. In the end, 11 articles were used, six for children and five for neonates.

To represent this process, the PRISMA 2020 methodology was used, and a flowchart was created, which was also provided by the developers of this research method.

After analyzing the articles, new studies were conducted to compare the improvement rates of patients based on vital sign categories, using parameters such as blood pressure, oxygen saturation, pain sensation level, respiratory rate, and heart rate. Data extracted from each study were organized into two separate Tables. The first group (Table 1) included the publication year, type of music therapy used, methodology analysis, and routinely used. The second set of Tables comprised data extracted from the articles, including systolic blood pressure, pain sensation level, respiratory rate, and heart rate.

**TABLE 1:**
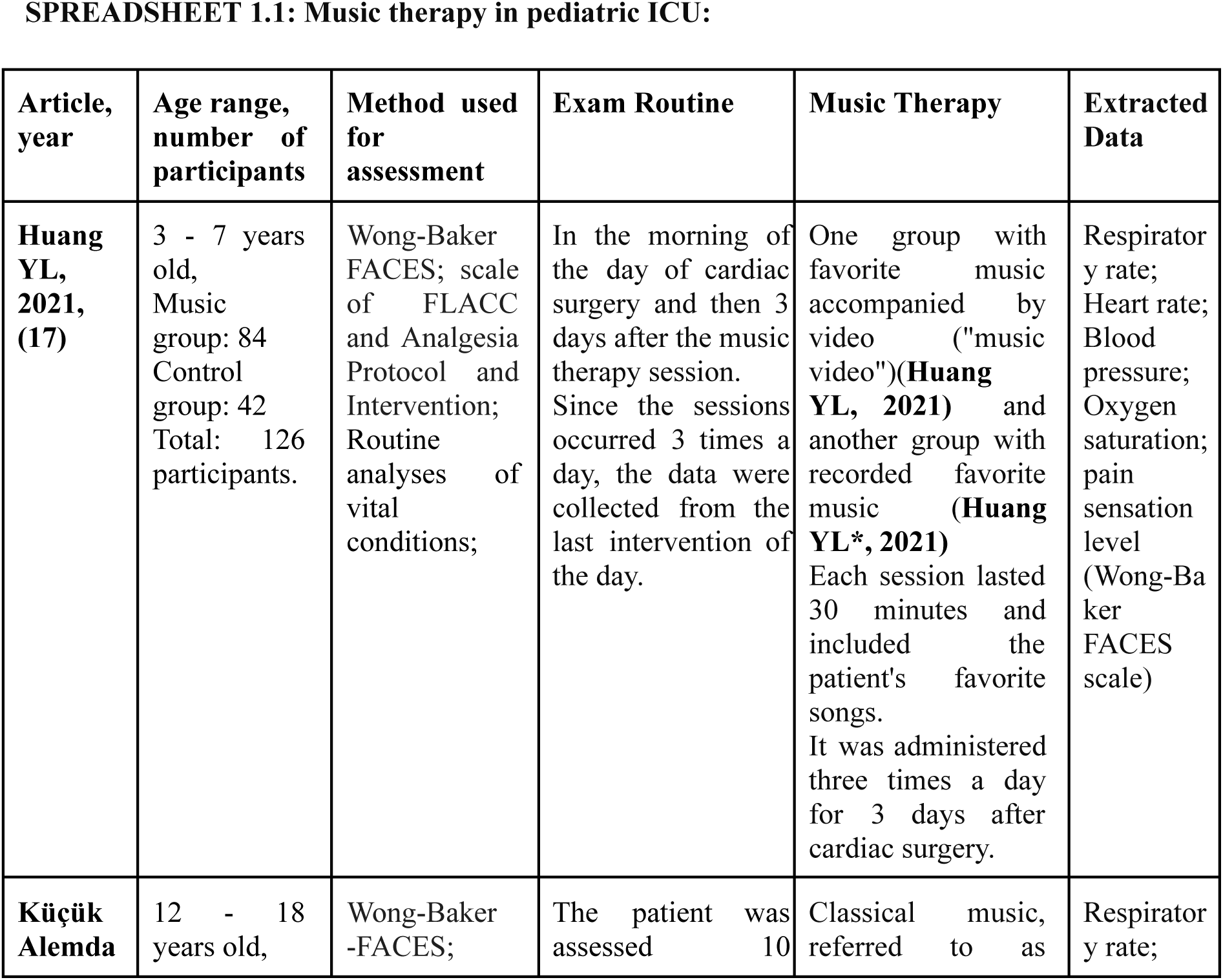
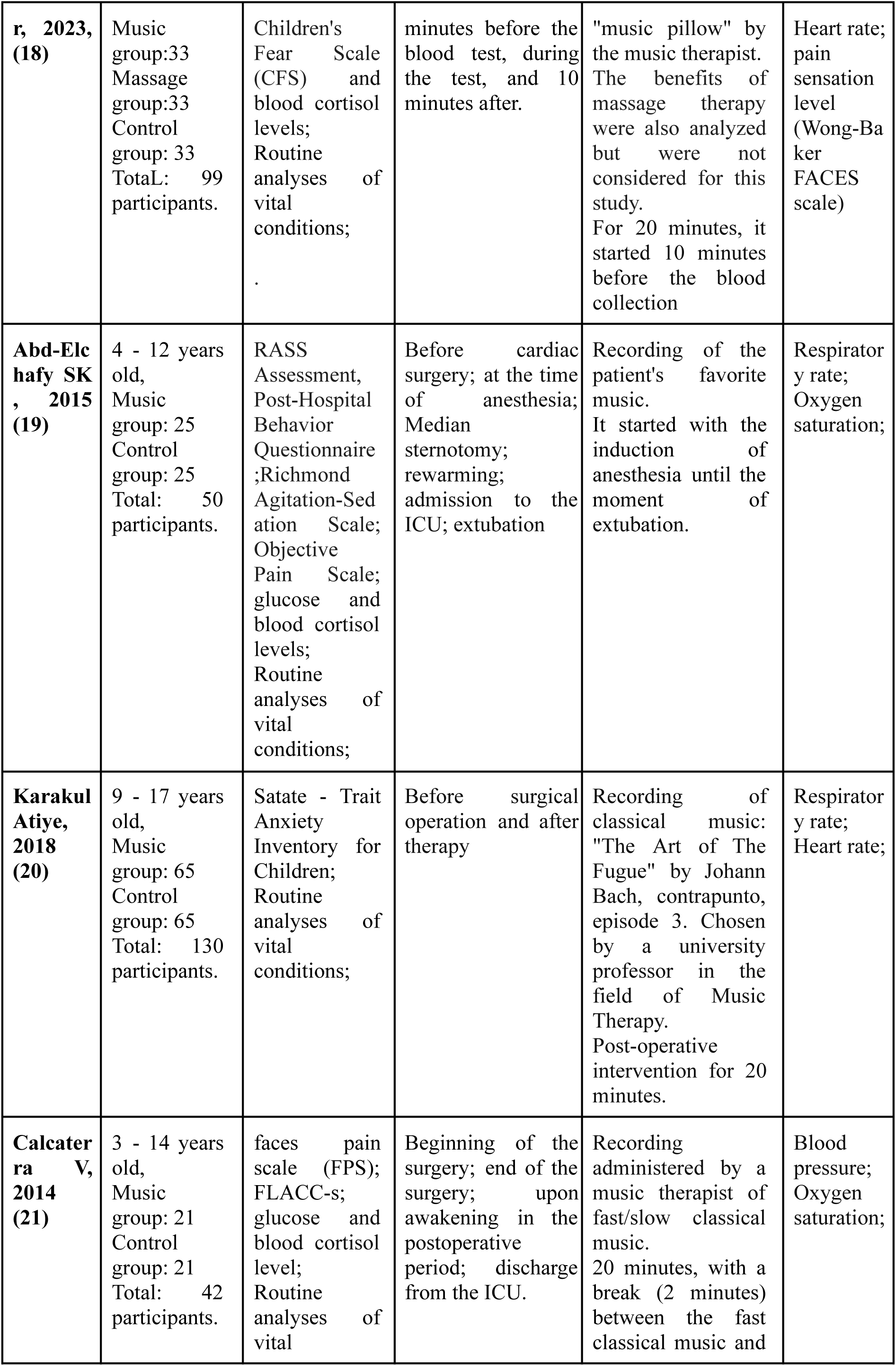
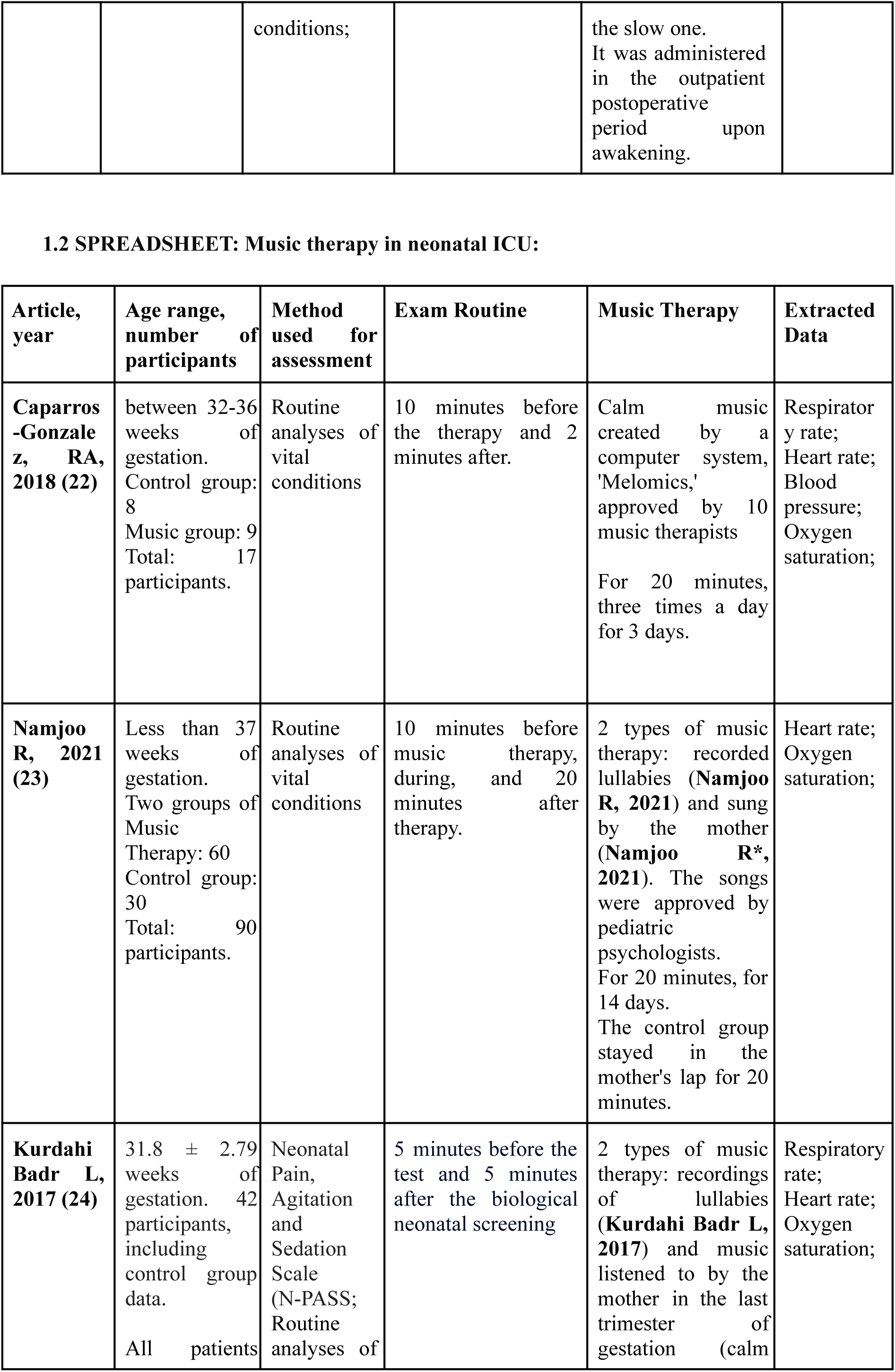
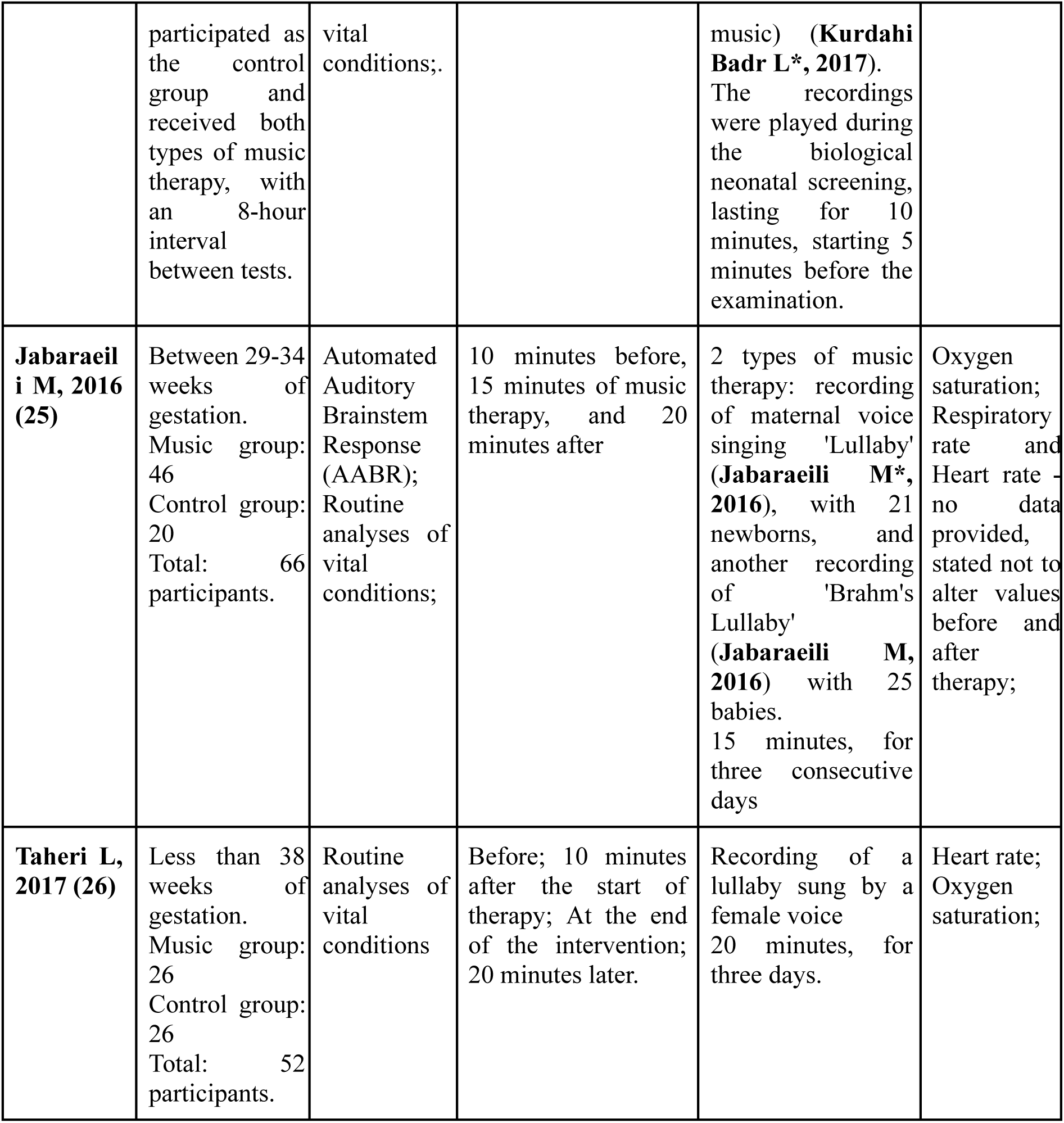
Spreadsheet with the main general information of the analyzed articles.

**TABLE 2:**
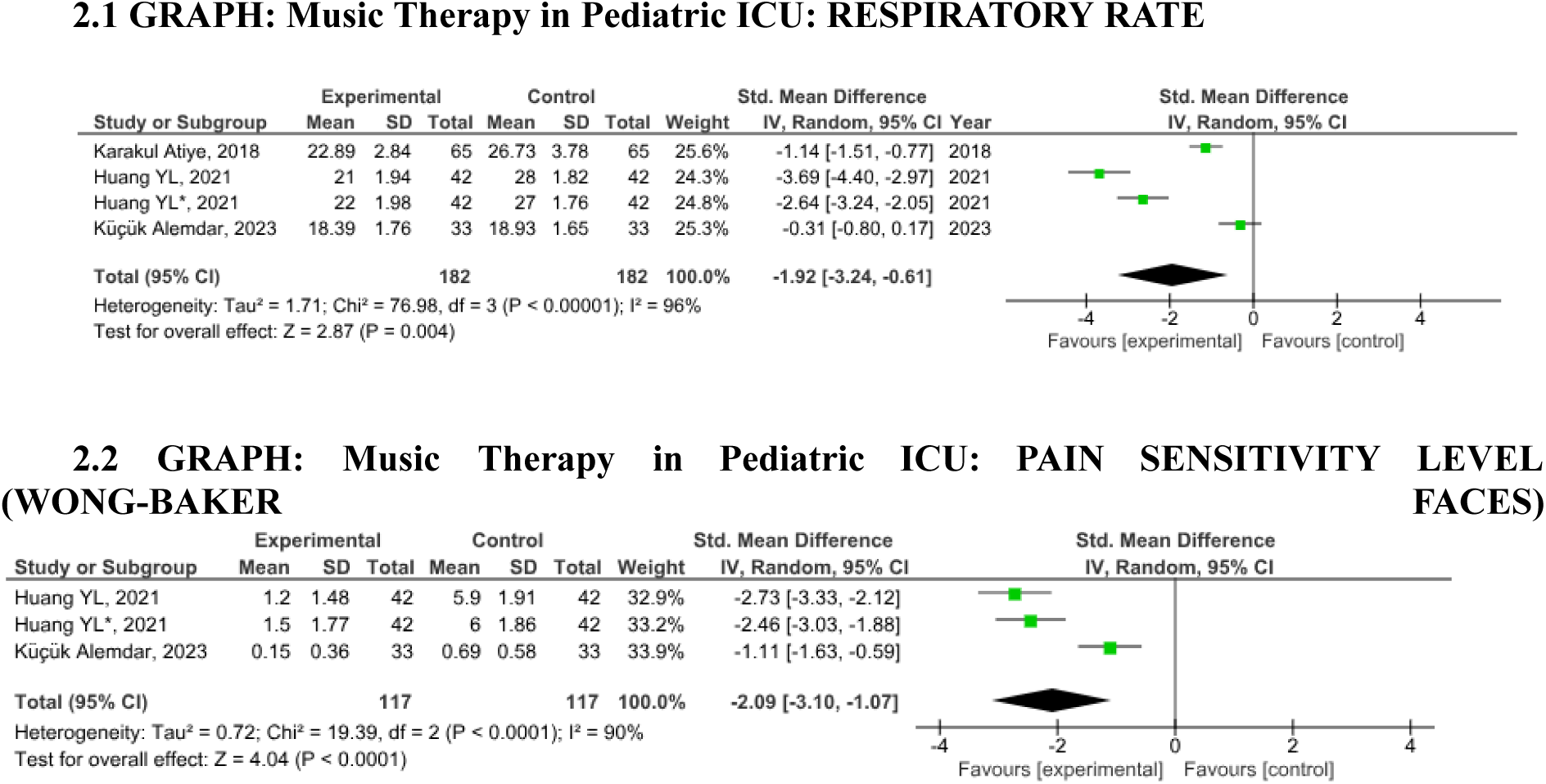
Forest Plot graphs, processed by the RevMan Software.

It is important to observe that the analysis of pain sensation level was only performed if multiple studies with the same analysis methodology were found.

As a result, a spreadsheet was compiled, along with the Forest Plot graphs, containing all the data before and after music therapy. Furthermore, specialized software, RevMan, which is available in the public domain, was employed for meta-analysis. Through RevMan, it was possible to analyze the heterogeneity index between the articles, confidence intervals, and p-values of each graph produced.

The values extracted from the chosen articles were characterized as continuous data since they were derived from the same population. Furthermore, it was assumed that changes in vital parameters could not be solely attributed to the use of music therapy, but could be influenced by other factors, such as the environment, the patient’s condition (routine ICU or before cardiac surgery), type of therapy, duration, and timing of vital sign measurement collection.

Furthermore, the most common and discussed vital parameters (respiratory rate, heart rate, blood pressure, oxygen saturation, and pain sensation level) among pediatric and neonatal articles were chosen for comparison. This was performed to compare the different outcomes between these populations of ICU patients.

In addition, an analysis comparing different types of music therapy among the studied age groups (pediatric and neonatal) was added to the study (Table 3). For this purpose, a series of graphs were created using Microsoft Excel to compare the simple means of vital values found in the articles before and after music therapy. It was not possible to conduct a meta-analysis for this purpose due to a lack of appropriate articles to analyze different types of music therapy, as filtered by the PRISMA 2020 method.

**TABLE 3:**
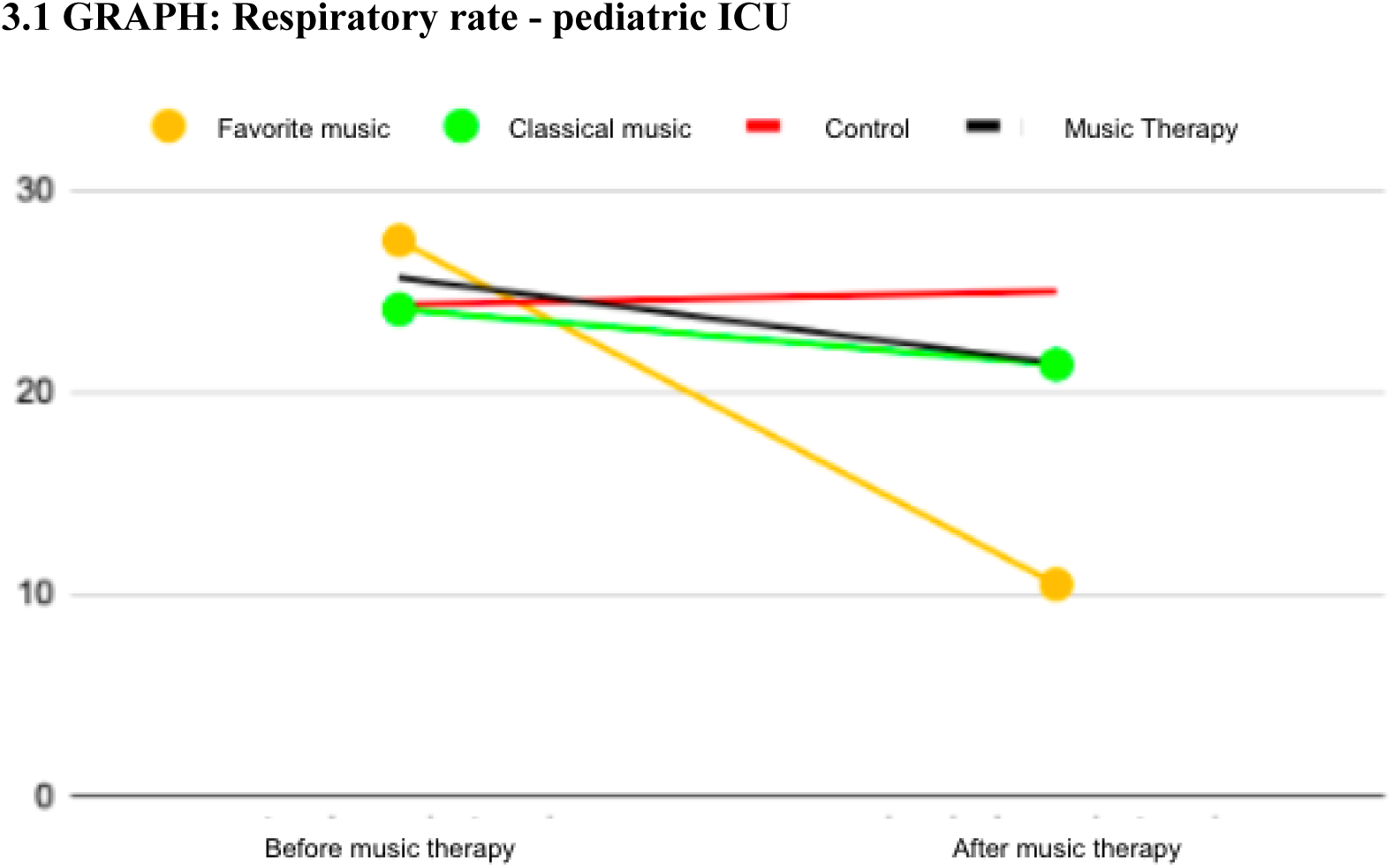
Average before and after graphs, conducted by the authors.

The different studied groups were groups that used classical music; music chosen by the patient; recorded or sung lullabies by the music therapist or mother; popular regional music with a calming effect; and electronic music created by a music therapist using a computer. In addition, there was a control group that did not receive the auditory stimulation of music therapy.

Finally, the selected texts were compared to parameters considered normal for each age group to heart and respiratory rates. For systolic blood pressure (14) The values were also compared with other articles published on these vital parameters (15).

### Assessing the Risk of Bias

All selected studies underwent manual analysis conducted by their authors to assess the risk of bias using the Rob.2 system developed by the company Concraine (16). Most of them was analyzed under the “randomized parallel trial” systematic; however, one of the studies [Kurdahi Badr L, 2021(24)] was of the “randomized crossover trial” type.

For studies analyzed under “randomized parallel trial”, aspects such as the randomization process; intended interventions; missing outcome data; measurement of the outcome; and selection of the reported result were evaluated.

While for the study with a “randomized crossover trial,” aspects such as the randomization process; intended interventions; missing outcome data; selection of the reported result; period, and carryover effects were considered.

The articles were manually classified according to the evaluated criteria using the flowcharts provided in the Rob.2 program.

Among the analyzed studies, eight were considered to be low risk and two with some risk. The studies considered to have some risk were one from the pediatric intensive care unit and the other from the neonatal sphere.

The analyzed studies were considered to be of low risk, and all were used to compose this work.

## Results

Initially, it was possible to analyze six articles focused on pediatric intensive care units by filtering the articles from the databases. One of these parameters was excluded during this work because it did not contain data on the previously chosen vital parameters. Additionally, it used a different methodology for quantifying anxiety levels, which makes it unsuitable for comparison on this type of scale. The Wong-Baker FACES scale was used to assess the patient’s pain sensation level.

In the neonatal intensive care unit sphere, five studies were found and analyzed.

They were therefore organized into a Table with important information for better understanding.

It was observed that all the articles used a passive methodology for music therapy, wherein the patient is the listener to the songs performed by music therapists, with the vast majority of the songs being recorded. In the active methodology, in which the patient is the musical agent, they can play music with professional health accompaniment, such as singing, playing, and doing improvised songs. However, this type of music therapy was excluded from this study due to the lack of published articles analyzing this type of music therapy and the initially established exclusion criteria (such as coherent information, and control group data)

Therefore, data on vital signs from each article were collected and organized into tables with pertinent information for conducting the meta-analysis, including the mean and standard deviation values before and after the intervention (Music Therapy). The same values were also considered for the control group (a group that did not receive music therapy stimulation).

With these Tables, it was possible to compare some of their values with the information found in the bibliography (14, 15). It is noticed that some are in the normal range for the patient’s age. However, others do not, and precisely these, the majority of the individuals analyzed, had some cardiac problems and were analyzed and exposed to Music Therapy during cardiac surgery. Therefore, it can be judged that this variation found was due to the vital conditions of the population analyzed.

Thus, their data was manually extracted from these formed Tables to compose the program of the RevMan software. In this way, it was possible to perform the meta-analysis based on vital parameters in comparison with the values before (control) and after the intervention (experimental). With this, these Forest Plot graphs were produced.

Therefore, the data on vital signs from each article were collected and organized into Tables with pertinent information for conducting the meta-analysis, including the mean and standard deviation values before and after the intervention (Music Therapy) The same values were also considered for the control group, which was not given music therapy stimulation.

With these Tables, it was possible to compare some of their values with information found in the bibliography (14, 15) It is noticed that some patients were in the normal range according to the patient’s age. However, others do not, and the majority of the individuals analyzed had some cardiac problems and were analyzed and exposed to Music Therapy during cardiac surgery. Therefore, it can be concluded that this variation was due to the vital conditions of the population analyzed.

It is observed that Music Therapy reduced the respiratory rate and pain sensitivity index in the pediatric ICU by 1.92 bpm (p-value: 0.004) and 2.09 Wong-Baker FACES units (p-value: <0.0001), respectively. Additionally, it increased oxygen saturation in pediatric and neonatal ICUs by 0.37 (p value:.0.001) and 0.2(p value: 0.04), respectively.

Most of the Graphs assessed that the data among the articles showed high or moderate values of heterogeneity (greater than 75%), with 4 being high, 3 moderate, and only 1 being 0%.

It is important to highlight that it was not possible to process data for the meta-analysis on blood pressure parameters in the neonatal ICU due to the lack of articles found. While in the pain sensitivity criterion, it was only possible to compare the articles with the same type of scale.

To analyze the type of Music Therapy, Microsoft Excel was used to compare the control group means of all music therapy articles according to the analyzed vital parameters and subgroups (types of interventions used). Analyzing the values before and after therapy.

The reason why it was not possible to perform a meta-analysis with these data was due to a series of factors, including a lack of articles to compare the subgroups with more than one single article - as some types had only one specimen to be compared; other methodologies were excluded due to the criteria established at the beginning of the research. Therefore, there was a massive loss of some articles, which made it difficult to perform a meta-analysis.

A significant portion of the Graphs showed a positive change in vital parameters with Music Therapy compared to control groups (patients who did not receive the same intervention), although not statistically proven using a meta-analysis. However, it is evident that among the analyzed subgroups, children and adolescents who listened to their favorite music performed better, as there was a greater positive variation between the values before and after, as well as in the neonatal ICU, with electronic music standing out - music created by a computer with the assistance of a music therapist.

## Discussion

Furthermore, most of the articles found used passive Music Therapy as the analysis method, in which the patient listens to melodies, sung or played by recording. Few articles explore other types of therapy, such as active therapy, in which the patient has access to musical instruments, performing, for example, improvisations.

One hypothesis for passive methodology being more present among articles studying vital parameters than active methodology is that it is more economical and practical to perform, as it can be administered by any healthcare professional, requiring only a sound resource. Additionally, not every patient may be able to perform the movements to use musical instruments, as this study analyzed the sphere of Intensive Care Units, where many children are in critical health conditions. As seen in this study (8), which compared the efficacy of active Music Therapy with passive, varying according to space, availability, and patient health - it was not used in the meta-analysis due to eligibility criteria.

One reason why passive methodology is more prevalent among articles studying vital parameters than active methodology is that it is more economical and practical to perform. It can be administered by any healthcare professional, requiring only a sound resource. Furthermore, not every patient may be able to perform the movements to use musical instruments, as this study examined the sphere of intensive care units, where many children are in critical health conditions. As seen in this study, which compared the efficacy of active music therapy with passive therapy, varying according to space, availability, and patient health; it was not used in the meta-analysis due to eligibility criteria.

There are certain controversies among the analyzed articles; some claim to reduce heart rate and not vary oxygen saturation (17,18,21) However, the meta-analysis found that the reduction found in these individual works was more likely to have been caused by other factors than Music Therapy, considering heart rate. The p-value was greater than 0.05. While oxygen saturation is statistically significant, as the p-value is less than 0.05, the variation is statistically significant. Nevertheless, clinically, as the increase in saturation was less than 0.5, numerous articles asserted that there was no distinction between the control and intervention groups.

Moreover, in Graphs where the p-value was less than 0.05, there was a high or moderate heterogeneity value. Although there were variations among the articles in the values found, due to the conditions the patients were in; the meta-analysis showed a statistically relevant value. Even with a variation in values among the articles used, possibly due to situational factors of each patient, there was still an improvement in the pain sensation index, heart rate, and oxygen saturation in the pediatric ICU. Therefore, demonstrating the impact of the intervention employed.

As for the other articles, which have a high heterogeneity index and high p-value, there are strong indications that Music Therapy did not directly interfere with these values. Due to the vital signs being so different among the articles and yet not showing a statistically valid difference between the values before and after the use of musical intervention. This proves that, possibly, the variation found was caused by other biological and situational factors.

It is worth highlighting that many of these patients were in different hospital situations, which may be a reason for presenting high heterogeneity values.

Regarding the technique, it is not possible to consider that it caused this variation among the articles, as the equipment used, and the healthcare team involved was similar among the articles. However, the exposure time of the patients to music therapy in each study was different, as well as the type of music presented, which may also confer differences between the values presented, causing heterogeneity.

Analyzing the moment of vital signs data collection among patients, it is noticed through the articles used that due to having different exposure times among them, this moment was also varied. On the other hand, for this study, only the values immediately before and after the use of the intervention were used to minimize the variation in vital signs of the studied individuals and cause any risk to the accuracy of the presented data.

Compared to other systematic review studies published during the period from 2013 to 2023; it is noticed that Music Therapy stands out for reducing pain and anxiety levels in its patients (27,28,29,30). There are still some studies highlighting the reduction in respiratory rate and increase in oxygen saturation (28,29,31). However, no article showed an improvement in blood pressure in Pediatric and Neonatal Intensive Care Units.

On the other hand, during the research, articles were found (34) denying the usefulness of music therapy, precisely because they have different models of using this type of therapy, which for these authors contributed to their conclusion that it is not possible to make a comparison of the applicability of music therapy compared to the control group.

However, through the statistical analysis performed, grouping all types of Music Therapy subgroups, it is possible to affirm that there is indeed a statistically significant difference between the values before and after the use of this type of intervention in the pediatric and neonatal population. Additionally, other factors can be evaluated, such as blood cortisol levels and duration of breastfeeding, which have good results in some articles and systematic reviews (29), as well as a reduction in postpartum depression rates (32,33) but were not analyzed throughout this work, as it was not possible to compare these rates with the pediatric population.

Few systematic reviews reported which subgroup of Music Therapy could be better. Among those found, only two articles highlighted classical and popular music (27,35), which aligns with the results found, as there was an emphasis on recording the patient’s favorite music in Pediatric Treatment Units, which can be classified as popular music.

Moreover, there are other studies (36,37) that examined the sphere of healthcare professionals and patientś parents. They claim to notice a significant reduction in anxiety levels. Many of them also noticed a more harmonious atmosphere between healthcare professionals and families. In one study (36), approximately 100% of respondents reported noticing an improvement in reducing stress levels. Thus, by lessening the stressful environment and fostering a more joyful, serene, and pain-free atmosphere; music therapy may help to lower the incidence of Post-Intensive Care Syndrome (PICS) in patients.

### Limitations

It is evident that due to the manual extraction and organization of data, some articles may have been missed, despite the use of various keywords for the search. Additionally, some analyzed articles had a wider age range than others, which could lead to variation in the overall means among vital parameters within the same article. The studies presented variations in the length of patient exposure to music therapy, which may have influenced the vital parameters of individuals exposed during the intervention.

### Conclusions

The requirement for additional articles in this field is evident, particularly in the sphere of active music therapy, to facilitate a more comprehensive and mathematical examination of diverse subgroups of music therapy. This type of therapy deserves recognition for its role in reducing patients’ pain sensitivity levels and significantly improving their quality of life, as they require fewer analgesics. It also promotes well-being in the pediatric and neonatal population. Despite periods of elevated stress, such as hospitalization in Intensive Care Units (ICUs) Furthermore, it was observed that the respiratory rate in the pediatric population experienced a decrease of approximately two breaths per minute and a slight increase in oxygen saturation levels. However, less positive results were obtained for the neonatal population, with only oxygen saturation showing improvement due to music therapy.

## Data Availability

All data produced are available online at the articles used during the research, and they are listed on the bibliographic section.

## Abbreviations and Acronyms

ICU: Intensive Care Unit

